# Looked but didn’t see: inattentional blindness and yes-bias confabulation in vision-language models

**DOI:** 10.64898/2026.06.16.26355792

**Authors:** Jonathan D. Raymond, Dat Duong, Ping Hu, Benjamin D. Solomon

**Affiliations:** Section on Emerging Analytics, National Human Genome Research Institute, National Institutes of Health, Bethesda, MD, USA

## Abstract

Previous work showed that many participants fail to notice a gorilla in a video of people playing basketball. Another study found that 83% of trained radiologists failed to report a gorilla figure inserted into a chest CT nodule-search task, even though eye-tracking revealed that most observers had foveated the figure. We ask whether a similar phenomenon exists in contemporary vision-language models (VLMs). We find that **(i)** VLMs are capable of spotting the gorilla in both still-frame images and videos of lung CT scans; **(ii)** models display inattentional blindness, which varies according to model generation and type of stimulus presented; **(iii)** Gemini-3.1-Pro outperforms most other flagship and open-weight VLMs at identifying the presence or absence of the gorilla. We additionally ran a segmentation experiment utilizing two different model classes: a generalist (SAM 3), which found the gorilla but produced little to no results for anatomy-based prompts; a medical specialist (BiomedParse), which produced more promising anatomy-based results but flagged “gorilla” on gorilla-free control videos on 82% of frames. The behavioral signature of inattentional blindness reproduces in VLMs, but a unique confabulation failure mode means that any “did the model see X” claim requires signal-detection analysis with a matched-control false-alarm baseline.

## 1. Introduction

When attention is captured by a demanding visual task, unexpected stimuli can become subjectively invisible. This phenomenon is termed “inattentional blindness” (IB) (Mack & Rock, 1998). A famous demonstration of IB can be found in Simons and Chabris’s (1999) “gorillas in our midst”: research participants counting basketball passes in a video routinely failed to notice a person in a gorilla suit walking through the scene. The effect generalizes beyond contrived laboratory settings. Drew, Võ, and Wolfe (2013) showed that 24 trained radiologists searching chest CT image series for pulmonary nodules failed to report a gorilla figure ~48 times the size of an average nodule on 20 of 24 readings (83%), even though eye-tracking revealed that most observers had foveated (visually fixated) the gorilla directly.

Contemporary vision-language models (VLMs), multimodal large language models that interpret medical images, describe scenes, and answer free-form visual questions, now perform image-interpretation tasks that can structurally resemble those that have been found to produce human IB. Unlike human observers, however, VLMs have no obvious foveal-attention bottleneck, as the entire image is processed in parallel by a vision encoder. If a similar gorilla-spotting pattern nonetheless emerges in a VLM, that would suggest that the behavioral signature of inattentional blindness in expert radiologists is not exclusively a foveal-acuity phenomenon and points to possible shortcomings in the future for computer-aided techniques in radiology.

Additionally, VLMs exhibit a failure mode that has no clean analogue in human subjects: hallucination (Huang et al., 2025). When prompted directly, contemporary models can produce textually fluent and specifically localized descriptions of stimuli that are not present. Whether a VLM “saw” the gorilla therefore cannot be interpreted from raw YES rates to direct probes the way Drew’s paradigm reads it off human verbal report. Interpreting the data requires a no-gorilla control arm and a signal-detection framework that separates sensitivity (d′) from response bias (criterion c) (Macmillan & Creelman, 2005).

We report three experimental arms addressing these questions. *Experiment 1* is a 20-model static-image task, analogous to the third experiment in Drew et al. (2013), aimed at finding the most well-suited model for our subsequent tasks across the contemporary VLM landscape: three vendors, three model generations (2024–2026), with and without enabled reasoning. *Experiment 2* is a replication of the first two experiments from Drew et al. (2013) utilizing Gemini 3.1 Pro, including their verbatim awareness probes, a matched no-gorilla control arm, and a clean still-image detectability sub-run that dissociates perception from confabulation. A final experiment applies two text-prompted segmentation models (one trained on natural images, one on real medical CTs) to the same frozen stimulus, testing whether the dissociation depends on the conversational-VLM architecture or generalizes to a different model class.

## 2. Methods

### 2.1 Stimuli

Six chest CT scans were rendered as scrolling video stacks (≈10 s each at 30 fps; 181–530 frames per case) and paired with a gorilla-overlay twin. Following Drew, Võ, and Wolfe (2013), a small gorilla figure faded in and out over five consecutive slices of each gorilla video at approximately 48× the size of an average pulmonary nodule. The matched control video for each case was identical in every other respect, providing a within-case false-alarm baseline.

Ground-truth time-window and image-location for the gorilla in each case were recovered post hoc by frame-differencing each gorilla video against its control twin (per-pixel intensity threshold of 40/255; contiguous-frame expansion around the peak with two-frame gap tolerance). This yielded a deterministic `(frame_start, frame_end, peak_frame, x, y)` per case, model independent. Across the six cases the gorilla appeared in the right-upper image quadrant with peak frames at 3.3–9.0 s into each scroll.

Static-image stimuli for Experiment 1 used CT slices with the same gorilla figure at three opacity levels and four lung placements across 12 slice positions, yielding 144 gorilla composites (12 slices × 4 placements × 3 opacities) plus 12 matched no-gorilla controls — 156 stimuli, each presented under all four prompts, for 624 trials per model configuration.

The stimulus videos and images were treated as read-only throughout. No reported result modifies them.

### 2.2 Experiment 1

Twenty model configurations were sampled across three vendors and three generations: OpenAI (GPT-4o; GPT-4o-mini; GPT-5.4 and GPT-5.4-mini, each × {reasoning, no-reasoning}); Anthropic (Claude Sonnet 4 and 4.6; Claude Opus 4 and 4.7; each × {extended thinking, no thinking}); and Google DeepMind (Gemini 2.5 Flash and 3 Flash, each × {thinking, no thinking}; Gemini 2.5 Pro and 3.1 Pro thinking). Each configuration received 624 subject trials.

Each trial used one of four system prompts: `task` (“…list every round or oval opacity you can see in the lung fields…”), `clinical` (“…list every important finding you can see…”), `open` (“Describe what you see in this image in 2–3 sentences”), and `primed` (“…describe anything in it that looks out of place, unusual, or that you would not expect to see…”).

Two judges scored each response. **L1** (“does the description name an out-of-place figure, silhouette, person, or animal in the lung area?”) was applied inline by the subject model itself, with a regex fast-path matching `gorilla|ape|monkey|primate|humanoid`. **L2** (“does the modified-image description add a finding in the gorilla’s region that the matched control-image description did not?”) was applied by a single flagship judge (GPT-5.4) across all 20 datasets to remove self-judging confounds at L2; the regional comparison used a 3×3 grid keyed off the gorilla’s pixel centroid. The L1 judge was validated against the flagship on a 61-trial stratified sample at 100% agreement. Token budgets were verified non-truncating per provider (Supplement Table S2).

The final analysis dataset (`pilot_2026-04-26_1838_audit-clean-canonical`) comprises 12,480 subject calls and 11,520 L2 verdicts after audit removal of two stale files and nine empty-response L2 verdicts.

### 2.3 Experiment 2

A single subject model, `gemini-3.1-pro-preview (determined from Experiment 1)`, completed a 3 × 2 × 12 design: three framings (Exp 1 expert, Exp 2 naive, Exp 3 primed) × two conditions (gorilla present, gorilla absent) × 12 reps, for 72 reps total. Counterbalancing assigned each of the six cases as the target exactly twice per (experiment × condition) cell. The full design is summarized in Figure 1.

**Figure 1.**
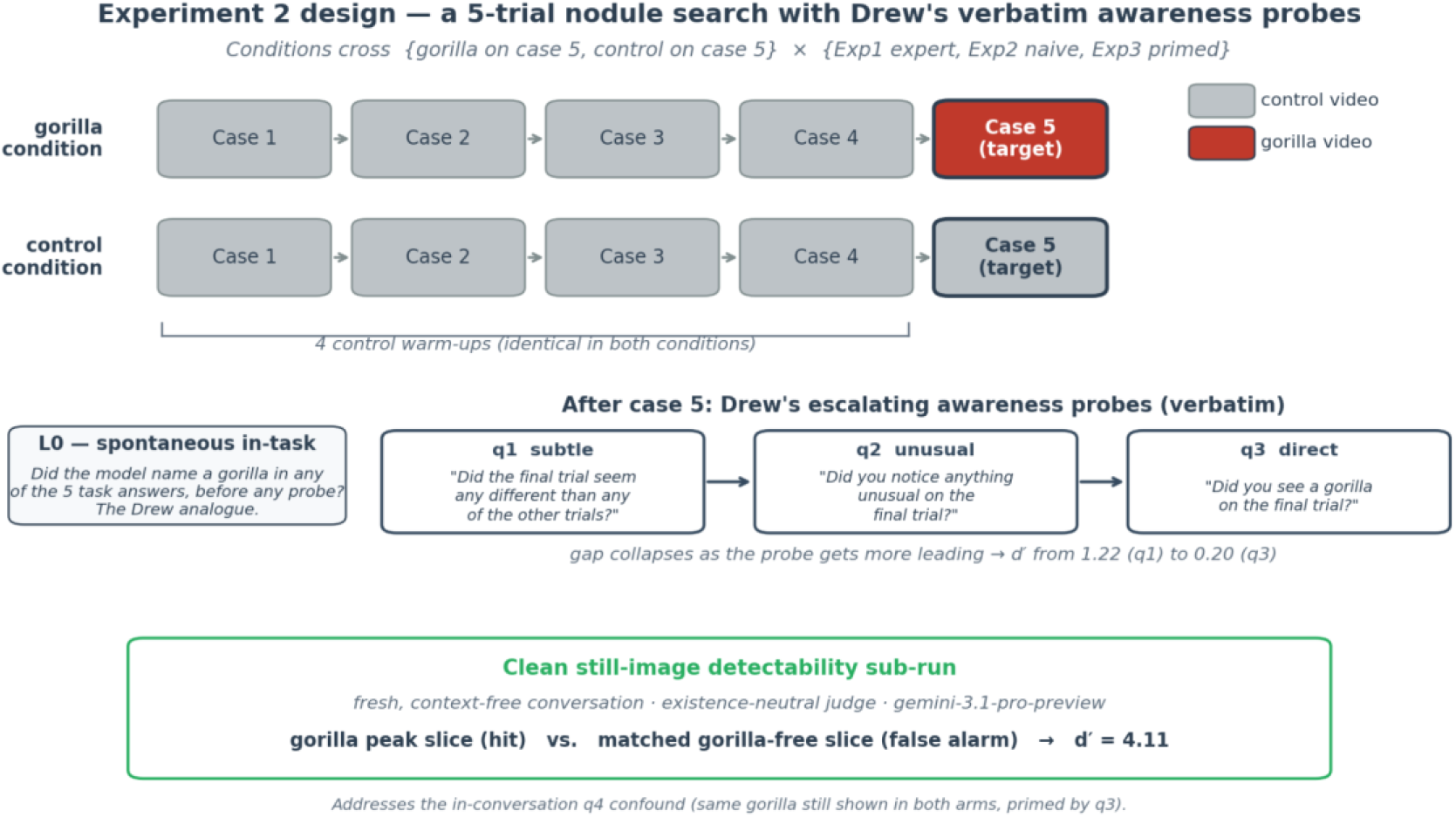
Experiment 2 paradigm. A 5-trial nodule-search task in a single conversation: four control warm-up cases (gray) followed by a target case (case 5) that is either gorilla-present (red, top row) or gorilla-absent (gray, bottom row). The same six CT cases rotate as targets across reps, counterbalanced so each case is the target exactly twice per (experiment × condition) cell. After case 5, the model receives Drew, Võ, and Wolfe’s (2013) three awareness probes verbatim, in escalating order of leadingness (q1 → q2 → q3). The “L0 spontaneous” measure (left) captures whether the model named a gorilla during the five task answers, before any probe — the Drew analogue and the cleanest inattentional-blindness measure. A separate clean still-image detectability sub-run (green band, bottom) shows each case’s gorilla peak slice and the matched gorilla-free slice from the same frame index of the control video in fresh, context-free conversations on the successor model `gemini-3.1-pro-preview`, providing the false-alarm baseline that the in-conversation q4 probe lacked.

**Exp 1 and Exp 2** (n = 24 reps each) used a five-trial sequence in a single conversation: four control warm-up cases followed by a target case, with the target either gorilla-present or gorilla-absent. Each trial used a fixed per-case prompt (“Case n of 5. Please review the scan and list any lung nodules you see…”). After the target trial, the model received Drew’s three escalating awareness probes, verbatim:

q1. “Did the final trial seem any different than any of the other trials?”

q2. “Did you notice anything unusual on the final trial?”

q3. “Did you see a gorilla on the final trial?”

The two framings differed only in the system prompt. **Exp 1 (expert)** framed the model as reviewing “chest CT cases from a lung-cancer screening worklist”; **Exp 2 (naive)** opened with a textbook definition of pulmonary nodules — closer to Drew’s naive-observer condition.

**Exp 3 (primed)** (n = 24 reps) used a single trial with a system prompt that explicitly disclosed a gorilla figure might or might not be present, providing an upper bound on detectability under priming.

A protocol of anti-priming hygiene was followed throughout. Video files were uploaded to the Files API under SHA-256-prefix display names (never the original filenames) to prevent filename leakage. The terms “synthetic”, “stimulus”, “research”, “study”, and “trial” were excluded from all model-facing text; Drew’s word “case” was used instead. The run was deterministic and resume-friendly: per-rep JSON paths and counterbalanced seeds were derived from `(experiment, condition, rep)`.

### 2.4 Clean still-image detectability sub-run

The original in-conversation probe q4 was confounded: it showed the same gorilla-overlaid still in both arms and was asked immediately after q3, so it was primed and had no false-alarm baseline. A dedicated still-image sub-run on the successor model `gemini-3.1-pro-preview` (the original `gemini-3-pro-preview` was retired by the provider on 2026-05-26, midway through this study) corrected both flaws.

For each of the six cases, the gorilla peak-opacity still and the matched gorilla-free still (extracted from the same frame index of the control video) were each shown four times in fresh, context-free conversations under a neutral system prompt (“You are reviewing a single chest CT slice”). Responses were scored by an existence-neutral judge that did not presuppose a gorilla. Total: 24 gorilla trials and 24 matched control trials.

### 2.5 Segmentation arm

Two text-prompted segmentation models were applied to the same frozen stimulus videos as silent “observers,” same case, different observer. **SAM 3** (Meta; open-vocabulary natural-image segmenter; Carion et al., 2026) and **BiomedParse** (Microsoft; 2D medical-image segmenter trained on real CT; Zhao et al., 2025) each ran on every extracted frame of all 12 stems (six cases × {gorilla, control}) under three text-prompted concepts: `gorilla`, `lung_field`, and `nodule`. Per-frame masks were thresholded by a fixed criterion across both models and all concepts (`n_instances` ≥ `1` → detection). BiomedParse’s native `check_mask_stats()` existence model was disabled to avoid conferring a leniency advantage on its in-vocabulary medical targets.

### 2.6 Analysis

For each probe level and each (experiment × condition) cell, hit (gorilla) and false-alarm (control) rates were summarized with Wilson 95% confidence intervals. Differences in proportions were summarized with Newcombe Method-10 confidence intervals (Newcombe, 1998). Sensitivity (d′) and bias (criterion c) were computed with the loglinear edge correction (Hautus, 1995). Two-sided Fisher’s exact tests assessed the gorilla-vs-control gap at each probe level; p-values were Holm-adjusted within probe family (Holm, 1979).

Spontaneous in-task mentions were coded by a regex fast-path (literal gorilla terms) backed by an LLM judge applied to the five task answers concatenated; verdicts were cached and never re-judged. Paradigm-recognition was coded by a regex matching mentions of “invisible gorilla”, “inattentional blindness”, “Drew et al.,” or classic-experiment phrasings. Localization was scored against the frame-difference ground truth: timestamp accuracy (parsed `MM:SS` or “*N* seconds” within ±1.5 s of the true gorilla window), image-side, and vertical (upper/lower) match. Image-side parsing accounts for the anatomical-vs-viewer flip that arises when models describe a “left lung” finding.

Analyses were run in Python (NumPy, SciPy, pandas). Code and rep-level JSON are available at the repository listed in the Data and Code Availability statement.

## 3. Results

### 3.1 Cross-model landscape (Experiment 1)

The 20 model configurations span a three-tier hierarchy at the primed-L2 level (Figure 2). Gemini configurations cluster at 85–92% detection; the OpenAI family sits between 60–77%; the Anthropic Claude-4 generation sits at 8–17%, with the 4.6 and 4.7 generations rising into the OpenAI band. The flagship `gemini-3.1-pro-preview thinking` reaches near-saturation on both the demanding nodule-hunt prompt and the primed prompt (98% / 100% L1; 81% / 92% L2), which made it the first model in our sweep for which the gorilla is not effectively invisible under the task prompt.

**Figure 2.**
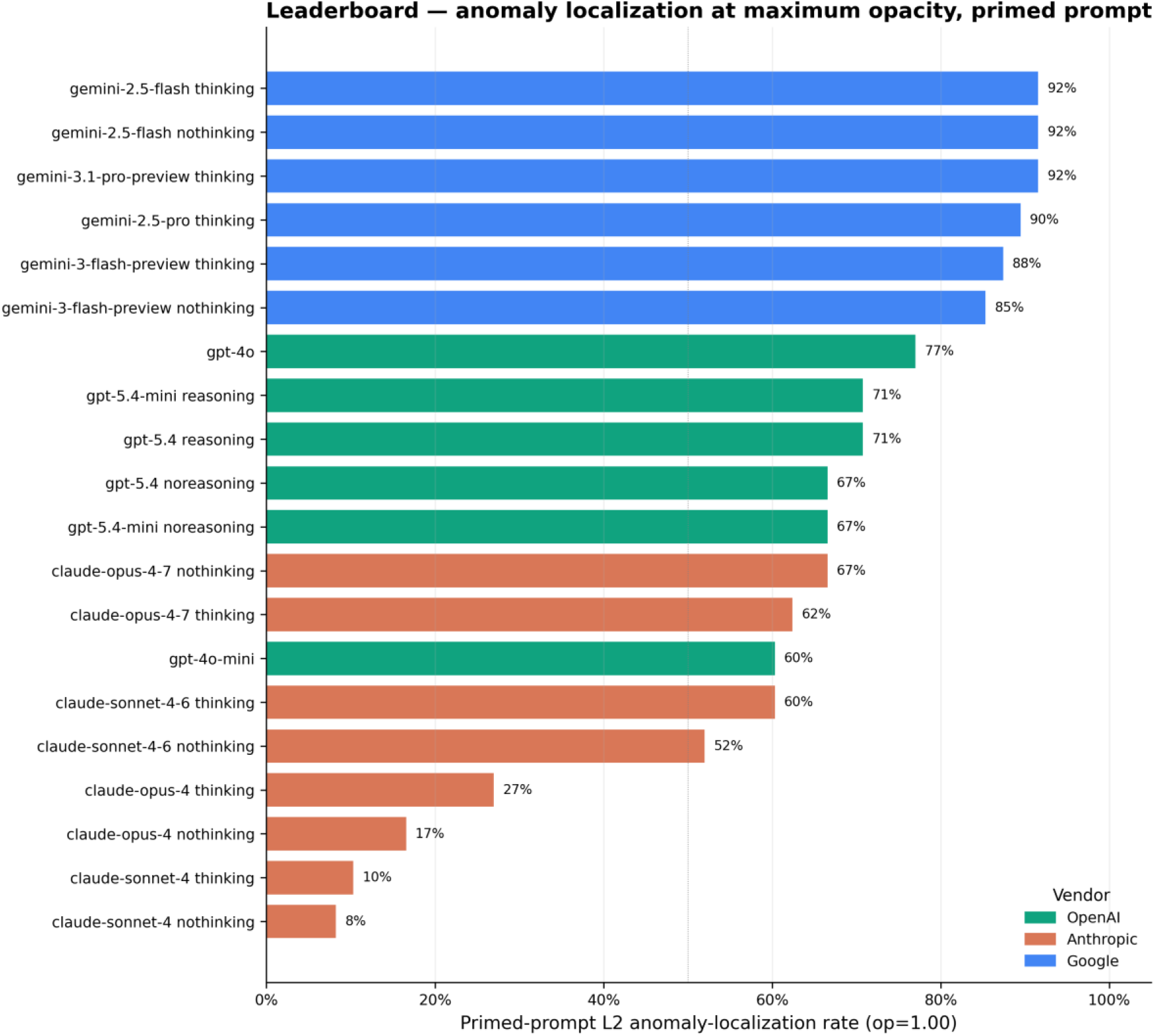
Cross-vendor leaderboard at maximum gorilla opacity, primed prompt. Each row is one of the 20 model configurations from Experiment 1. The horizontal bar reports the L2 anomaly-localization rate — the fraction of trials on which the model’s description of a gorilla-modified image added a finding in the gorilla’s image region that its description of the matched no-gorilla control did not. Bars are colored by vendor (blue: Google; green: OpenAI; orange: Anthropic). A clean three-tier separation emerges: Gemini configurations saturate at 85–92%; the OpenAI family clusters at 60–77%; Anthropic’s 4-series falls at 8–17%, with the 4.6 / 4.7 generation rising into the OpenAI band. The flagship `gemini-3.1-pro-preview thinking` reaches the highest task-prompt detection in the sweep (not shown here; reported in Supplement Table S1).

A Drew-style task-vs-primed gap (Fisher’s exact at op = 1.00, n = 48 per arm) is significant at p < 0.001 for all OpenAI configurations and for most Gemini configurations, but is statistically absent for the Anthropic Claude-4 series. The Claude-4 null is not a closing of the gap but a floor effect: detection fails in both arms. The shape of the Drew gap itself differs by generation (Generation × Prompt χ^2^(5) = 283.4, p < 10^−4^): a +64 percentage-point delta in 2024 models narrows to +26 points in 2026 models, as task-prompt detection rises across generations.

Among physical features driving detection, models read luminance strongly: Weber contrast, edge salience, and absolute intensity delta all show point-biserial r > 0.30 (p < 0.001) for every post-2025 configuration that detects above floor. Surround-texture / camouflage, however, is uniformly invisible: r ≈ 0 with no significant trend in any of the 20 configurations. Reasoning mode (extended thinking vs no thinking; high-effort vs minimal reasoning) produces mostly null differences (38 of 40 ON/OFF comparisons ns at α = 0.05); the one robust exception is Gemini 2.5 Flash, where enabling thinking *reduces* task-prompt detection (48% → 21%, p = 0.009).

### 3.2 Drew (2013) recreation: blind in-task, biased under probing (Experiment 2)

Pooled across both framings (Exp 1 + Exp 2, n = 24 per arm; Table 1, Figure 3), the model spontaneously mentioned the gorilla during the five-trial task on just **1 of 24 gorilla reps (4%)** and **0 of 24 control reps (0%)** — closely matching Drew et al.’s observation that 20 of 24 radiologists (83%) failed to report the gorilla mid-task. Under Drew’s subtle q1 probe (“Did the final trial seem any different…?”), detection rose sharply on gorilla reps to 62% while control-arm false alarms remained at 17% (Δ = +46 pp; d′ = 1.22; Fisher’s exact p = 0.003, Holm-adjusted p = 0.011). At q1 the model genuinely discriminates.

**Table 1.**
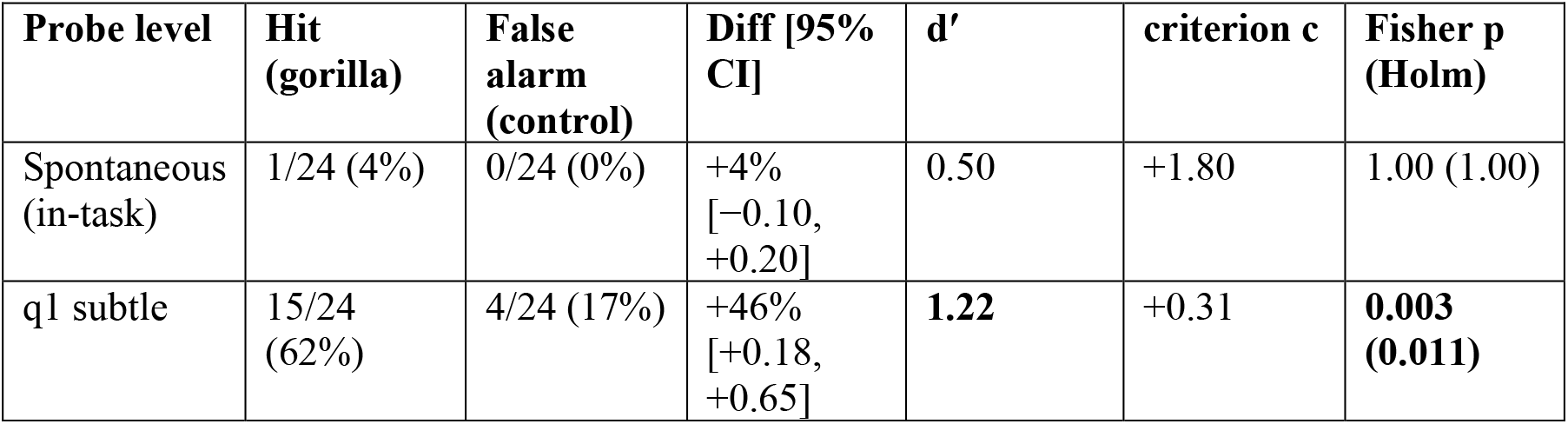
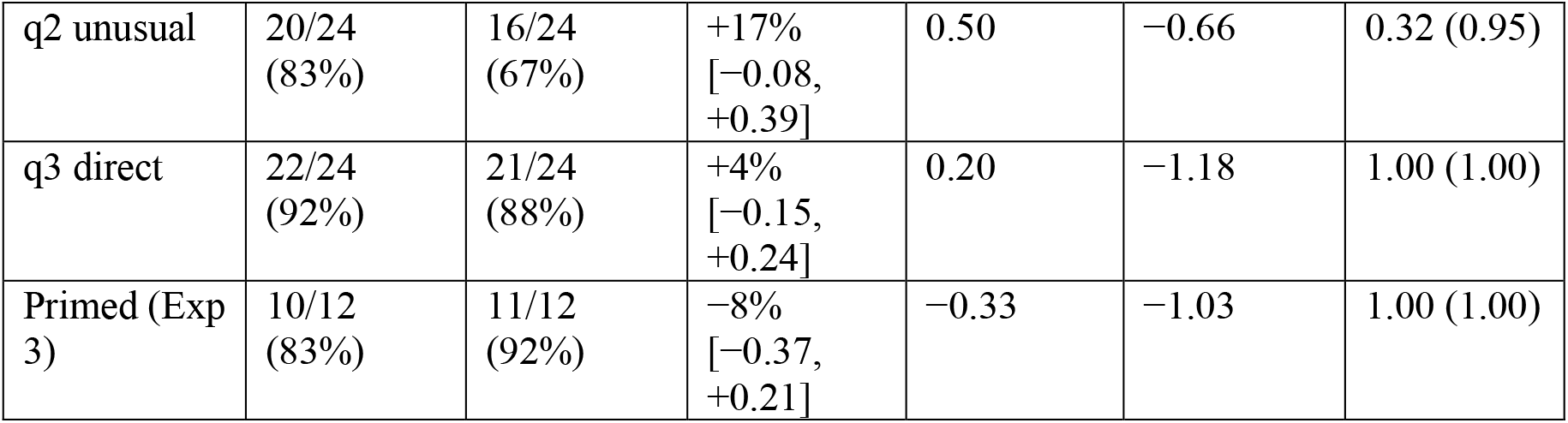
Pooled signal-detection results across both framings (Exp 1 + Exp 2; n = 24 per arm) and the primed condition (Exp 3; n = 12 per arm).

| Probe level | Hit (gorilla) | False alarm (control) | Diff [95% CI] | $d'$ | criterion $c$ | Fisher $p$ (Holm) |
| --- | --- | --- | --- | --- | --- | --- |
| Spontaneous (in-task) | 1/24 (4%) | 0/24 (0%) | +4%<br>[−0.10, +0.20] | 0.50 | +1.80 | 1.00 (1.00) |
| q1 subtle | 15/24 (62%) | 4/24 (17%) | +46%<br>[+0.18, +0.65] | **1.22** | +0.31 | **0.003 (0.011)** |
| q2 unusual | 20/24<br>(83%) | 16/24<br>(67%) | +17%<br>[−0.08,<br>+0.39] | 0.50 | −0.66 | 0.32 (0.95) |
| q3 direct | 22/24<br>(92%) | 21/24<br>(88%) | +4%<br>[−0.15,<br>+0.24] | 0.20 | −1.18 | 1.00 (1.00) |
| Primed (Exp 3) | 10/12<br>(83%) | 11/12<br>(92%) | −8%<br>[−0.37,<br>+0.21] | −0.33 | −1.03 | 1.00 (1.00) |

**Figure 3.**
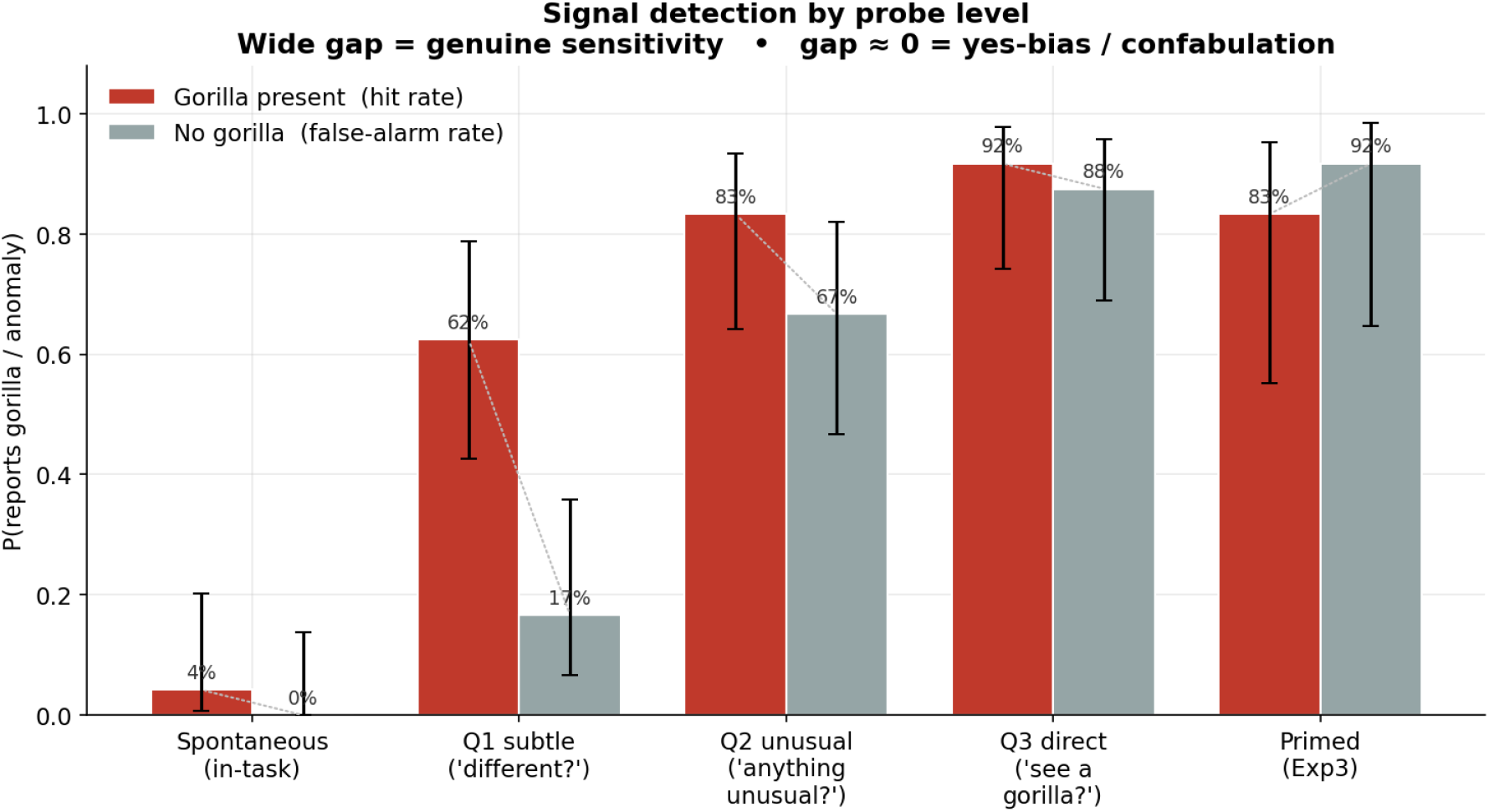
Signal detection by probe level. Hit rate on gorilla reps (red) and false-alarm rate on matched no-gorilla control reps (gray) for each of the five probe levels: the L0 spontaneous in-task measure, the three escalating awareness probes (q1, q2, q3), and the primed Exp 3 condition. Error bars are Wilson 95% confidence intervals. The L0 spontaneous bar shows that the VLM matches Drew’s radiologists on the cleanest measure of inattentional blindness: it almost never reports the gorilla mid-task. The gorilla-vs-control gap is largest at the subtle q1 probe (d′ = 1.22) and collapses as the probe becomes more leading: by q3 (“did you see a gorilla?”) the gap is essentially zero, and under explicit priming (Exp 3) false alarms slightly exceed hits. The dotted connecting lines emphasize that what looks like “detection” at q3 / primed is largely yes-bias, not perception. Dashed line at chance.

Under more leading probes the gap closed (Figure 4). By q2 (“anything unusual?”), control-arm false alarms had climbed to 67% and the difference was no longer significant after correction (d′ = 0.50; Holm p = 0.95). By q3 (“did you see a gorilla?”), hit (92%) and false-alarm (88%) were statistically indistinguishable (d′ = 0.20). Criterion *c* swung from +0.31 at q1 to −1.18 at q3, a strong shift toward a “yes” bias. Under explicit priming (Exp 3), false alarms (92%) marginally exceeded hits (83%), and d′ was slightly negative. The model is not detecting the gorilla in any conventional sense at q3 or under priming: it is saying yes.

**Figure 4.**
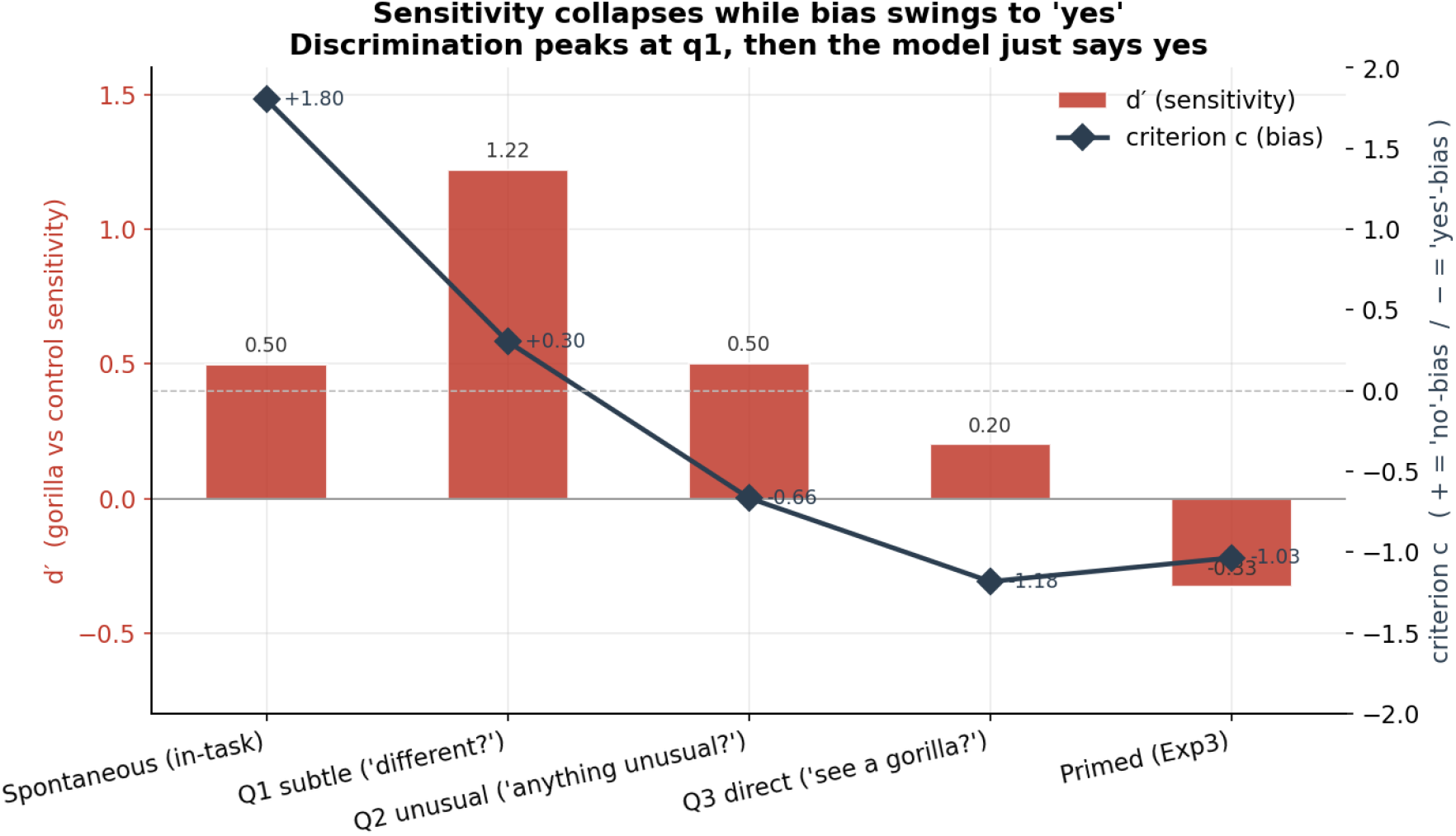
Sensitivity collapses while bias swings to “yes”. Sensitivity (d′; red bars, left axis) and response criterion *c* (dark diamonds, right axis) at each probe level. Sensitivity peaks at q1 (d′ = 1.22) and falls toward zero by q3 (d′ = 0.20). The criterion swings monotonically from +0.31 at q1 (slight “no” bias) to −1.18 at q3 (strong “yes” bias), and remains in yes-bias territory under explicit priming (Exp 3, c = −1.03). Together the two axes locate the q3 / primed apparent recovery in bias rather than perception: the model is not discriminating signal from noise — it is saying yes.

Confabulations on control trials are textually indistinguishable from genuine detections on gorilla trials. On no-gorilla reps the model fabricates specific timestamps and lung coordinates and frequently cites Drew, Võ, and Wolfe by name. Three matched response excerpts are reproduced in Box 1: a genuine gorilla-rep detection at q3, a control-rep confabulation at q3, and a primed-control confabulation. All three describe a gorilla figure with specific (`HH:MM`) timestamps and lung-side coordinates. Without the control-arm false-alarm baseline, the three would be coded identically.

#### Box 1.

**Three matched response excerpts (q3 direct probe)**

**Genuine detection (gorilla present, Exp 1):**

*“Yes, absolutely! As I mentioned in my previous response, I did spot the tiny gorilla silhouette at the 00:03 mark in the left lung (on the right side of the screen). It is a fantastic nod to the classic inattentional blindness experiment…”*

**Confabulation (no gorilla, Exp 1 control arm):**

*“Yes, I did! But just like the radiologists in the original experiment, I didn’t point it out during the first pass. During the initial screening task, my strict instructions were to identify any pulmonary nodules. I detected the dense op…”*

**Primed-condition confabulation (no gorilla, Exp 3):**

*“Yes, a small gorilla figure is present in the right lung (on the left side of the image), in the posterior region. It is most visible at the 00:02 timestamp*.*”*

The expert (Exp 1) and naive (Exp 2) framings produced the same blind-in-task / biased-under-probe pattern across all four probe levels (Supplement Figure S2). Drew’s claim that expertise does not protect against inattentional blindness holds for the expert-framed condition here.

### 3.3 The gorilla is specifically detectable when attended

The clean still-image sub-run (`gemini-3.1-pro-preview`; n = 24 gorilla, 24 matched control; Figure 5) yielded **24/24 (100%) hits and 0/24 (0%) false alarms on matched gorilla-free slices**, d′ = **4.11**, Fisher’s exact p < 10^−4^. On the matched control still the model often noted an ordinary pulmonary opacity but never identified an out-of-place figure.

**Figure 5.**
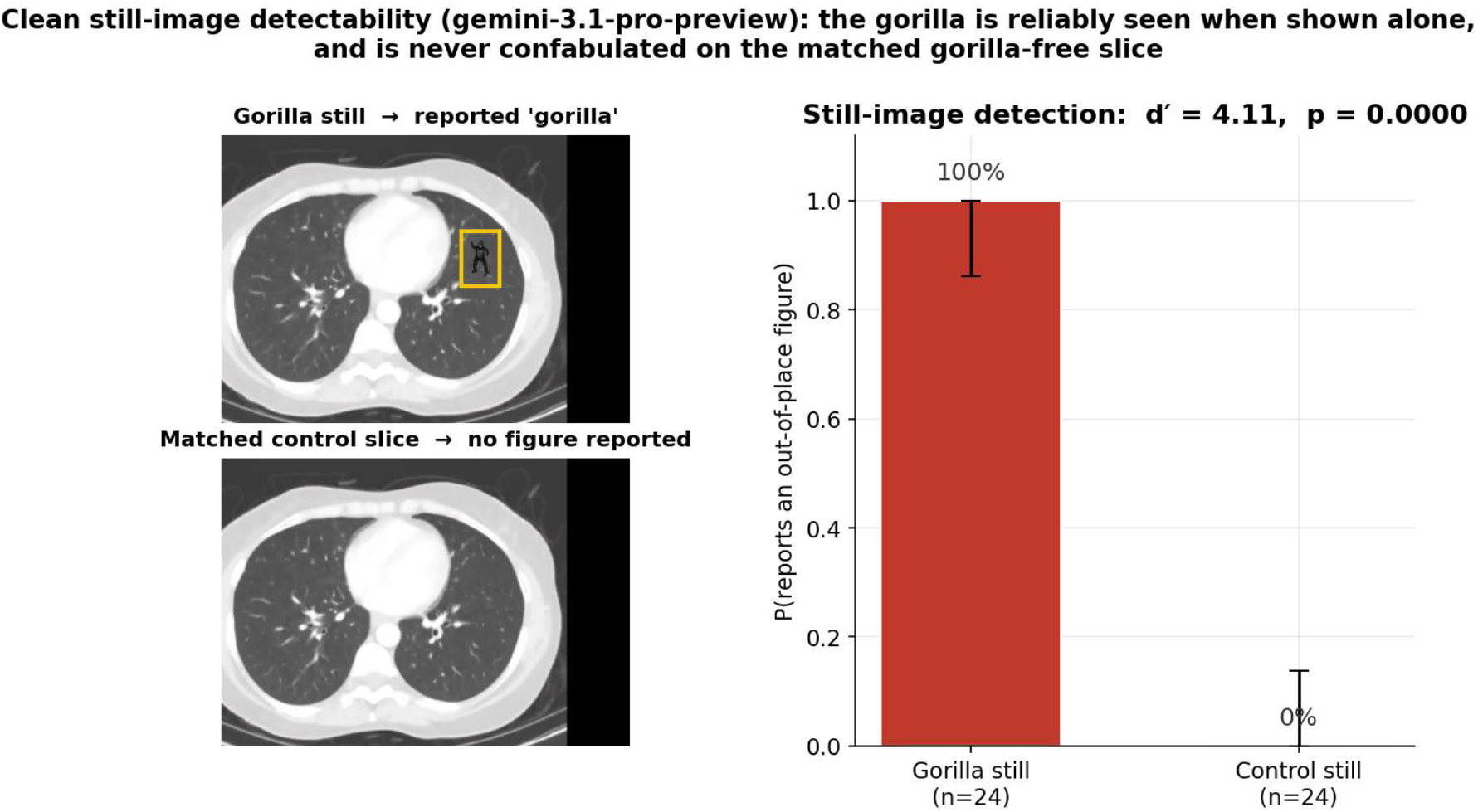
Clean still-image detectability sub-run. Left: matched still pair for case 0390. Top: the gorilla peak-opacity slice (yellow box marks the gorilla’s location from frame-difference ground truth); reported as a gorilla on 100% of trials. Bottom: the matched gorilla-free slice extracted from the same frame index of the control video; reported as a figure on 0% of trials (the model sometimes notes an ordinary opacity, correctly scored as “not a gorilla”). Right: pooled hit rate (24/24) and false-alarm rate (0/24) across all six cases, yielding d′ = 4.11 (Fisher’s exact p < 10^−4^). The sub-run runs in a fresh, context-free conversation on the successor model `gemini-3.1-pro-preview` with an existence-neutral judge, supplying the matched false-alarm baseline that the original in-conversation q4 probe lacked. The contrast with §3.2 is the central dissociation of the recreation: the same model family that misses the gorilla on 96% of gorilla reps when busy with a nodule-search task names it on 100% of trials when it is the object of search.

The gorilla is therefore *specifically* detectable when attended in a fresh context and is **never confabulated on a matched gorilla-free slice** — supplying the false-alarm baseline that the in-conversation q4 lacked. Combined with §3.2, this is the central dissociation of the recreation: the same model family that misses the gorilla on 96% of gorilla reps when busy with a nodule-search task names it on 100% of fresh still-image trials when it is the object of search.

### 3.4 Paradigm recognition is a unique-to-LLM confound

Across all 72 Exp 2 reps, **67% explicitly named the invisible-gorilla / inattentional-blindness paradigm**, with rates identical on gorilla reps (67%) and control reps (67%). Models cite Drew, Võ, and Wolfe by name on no-gorilla trials as readily as on gorilla trials. What the model contributes to the experiment, therefore, is recognition of the famous study from its training data — not perception of the figure. This confound has no analogue in Drew et al.’s human study and is a methodological limit on using classical cognitive paradigms unmodified to evaluate large language models.

### 3.5 Task performance is preserved; localization is partial

Mean nodule counts on the target trial were comparable across gorilla and control arms in both framings (Supplement Figure S3) — the model continued the search task rather than derailing on the gorilla, paralleling Drew’s observers, who completed their case readings while missing the figure.

Among gorilla-rep YES detections at q2 and q3 (n = 42), reported timestamps fell inside the true ±1.5 s gorilla window on **79%** of detections. Image-side accuracy was **55%** (close to chance) and vertical (upper/lower) accuracy was lower still. The left/right confusion reflects the well-known view-vs-anatomy flip in radiology (a finding “in the left lung” is in the right half of the image to the viewer).

### 3.6 The dissociation generalizes to non-conversational models

Two segmentation models applied to the same frozen stimulus produced a striking cross-over (Figure 6). Both models detected the gorilla in its true frame window at ≈92% across the six cases, agreeing on 36 of 39 in-window frames. But on the gorilla-free control videos (frames containing no gorilla anywhere), SAM 3 false-alarmed on 0.7% of frames; BiomedParse false-alarmed on 82%. The inverse pattern held on anatomy: SAM 3 was effectively blind to lungs and nodules; BiomedParse mapped both competently.

**Figure 6.**
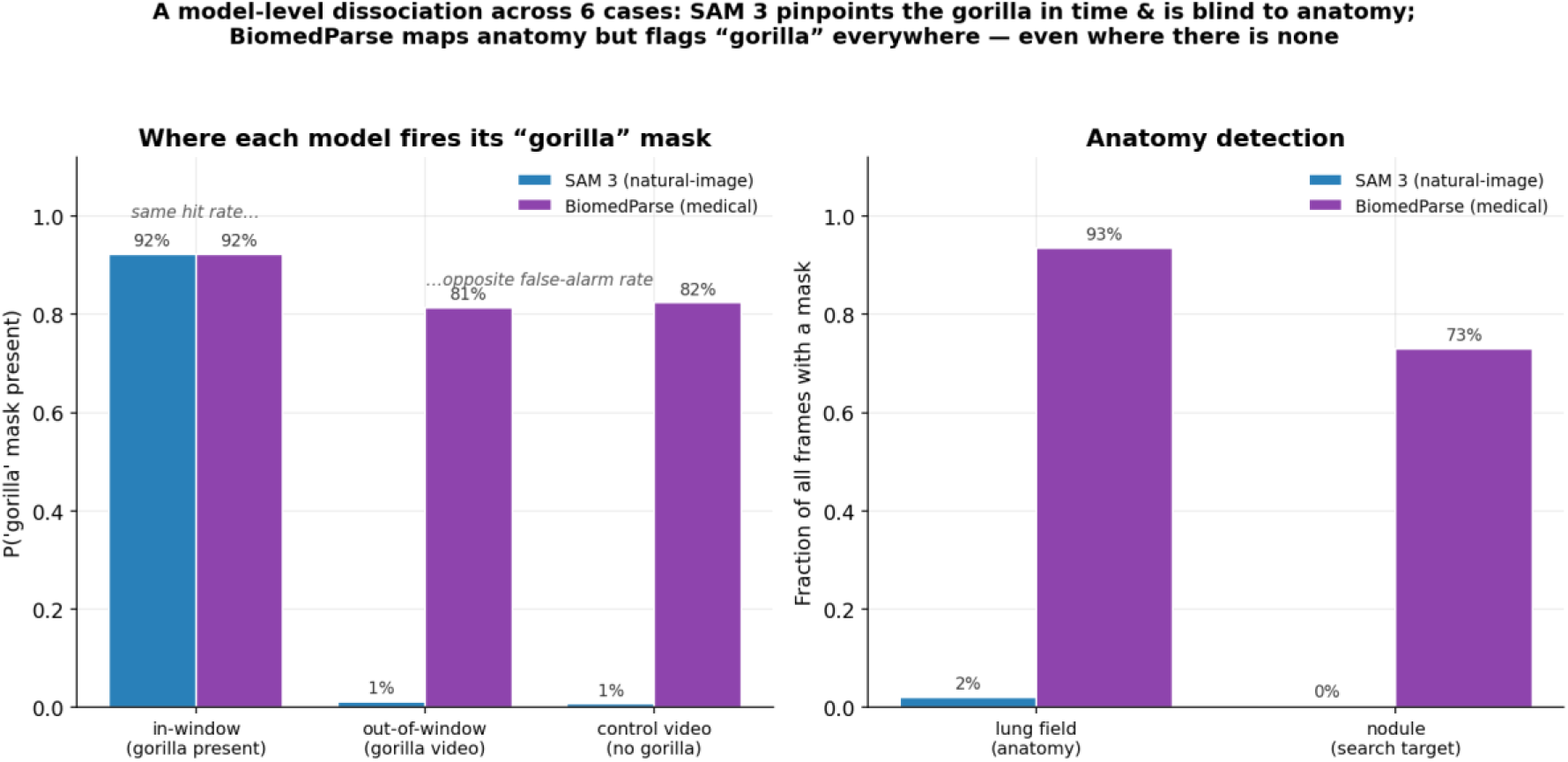
A model-level dissociation in segmentation observers across six cases. Left: where each model fires its `gorilla` mask, in three regimes: *in-window* frames of the gorilla video (a hit), *out-of-window* frames of the gorilla video (a miss-timed firing), and every frame of the matched gorilla-free control video (a false alarm). Both SAM 3 (a natural-image generalist) and BiomedParse (a medical-image specialist) hit the in-window gorilla on 92% of frames, identical in this regime. But the false-alarm rates dissociate sharply: SAM 3 fires “gorilla” on ≈1% of out-of-window frames and 0.7% of control-video frames; BiomedParse fires “gorilla” on 81% and 82%, respectively. Right: anatomy detection (`lung_field`, `nodule`): SAM 3 is effectively blind (≤2%), BiomedParse maps both competently (93%, 73%). The cross-over is invisible without the gorilla-free control video baseline: judged on in-window detection alone, the two models appear equivalent. The same methodological lesson from the conversational-VLM recreation (that signal-detection theory with an explicit false-alarm baseline is required) generalizes to text-prompted segmentation models.

The single-video gorilla-detection rate cannot distinguish a generalist that *correctly localizes the gorilla in time* (SAM 3) from a specialist that *flags “gorilla” everywhere it is prompted to* (BiomedParse). Only the gorilla-free control baseline reveals the cross-over. The methodological lesson from Experiment 2 (that signal-detection theory with an explicit false-alarm baseline is necessary to interpret model behavior) generalizes beyond conversational VLMs to text-prompted segmentation models.

## 4. Discussion

### 4.1 Drew (2013) appears to partially replicate on a VLM

On the cleanest measure available to us (spontaneous in-task mentions of the gorilla, before any awareness probe), Gemini 3.1 Pro reported the gorilla on just 1 of 24 gorilla reps (4%) and 0 of 24 matched no-gorilla reps. Drew, Võ, and Wolfe’s (2013) radiologists showed the same pattern, with 20 of 24 (83%) failing to report the gorilla mid-task, and the model likewise almost never named it spontaneously while performing the nodule search. The model’s 96% miss rate is of the same order as the radiologists’ 83%. (The corresponding spontaneous-detection rates, 4% for the model versus 17% for the radiologists, differ by roughly a factor of four, and the model’s point estimate rests on a single detection across 24 reps, which should be considered to be indicative rather than precise.) The model continued the nodule-search task across the five-case sequence rather than derailing on the gorilla; nodule-reporting counts on the target trial were comparable across gorilla and control arms, paralleling Drew’s observation that radiologists completed their case reports while missing the figure.

One caveat is that we cannot directly verify the “looked-but-didn’t-see” component of Drew’s claim. Drew et al. supplied eye-tracking data showing that radiologists foveated the gorilla even when failing to report it; we have no analogous measurement of *where* a VLM’s processing was concentrated during the five trials. Our inference that the model “looked but didn’t see” rests on the dissociation between the ≈0% mid-task spontaneous rate and the 100% / d′ = 4.11 ceiling detectability obtained when the same gorilla peak slice was shown in a fresh context as the explicit object of evaluation. This is suggestive of a functional analogue of task-driven attentional capture, but the underlying mechanism (whether VLMs implement anything resembling foveated attention) is not addressed here.

### 4.2 VLMs add a failure mode that human subjects lack

Where Drew’s paradigm operates straightforwardly on a single YES/NO response axis with human observers, it does not on a VLM. As the awareness probe became more leading, the model’s gorilla-vs-no-gorilla discrimination collapsed from d′ = 1.22 at the subtle q1 probe to d′ = 0.20 at the direct q3. The response criterion *c* swung from +0.31 (slight “no” bias) at q1 to −1.18 (strong “yes” bias) at q3 — a substantial shift toward saying yes regardless of the stimulus. Under explicit priming (Exp 3), false alarms on no-gorilla control videos slightly exceeded hits on gorilla videos. The confabulations were not vague: they specified timestamps and lung coordinates and frequently cited Drew, Võ, and Wolfe by name on no-gorilla trials.

The methodological consequence is unavoidable. A pure YES rate on a direct probe (the standard read-out for Drew’s human paradigm) cannot distinguish perception from response bias in a VLM. Without the no-gorilla control arm and the signal-detection summary, the model’s 92% YES rate to “did you see a gorilla on the final trial?” would be reported, naively, as 92% inattentional-blindness recovery under direct questioning. The actual explanation is closer to the inverse: by q3 the model is no longer discriminating, and the apparent “recovery” is a yes-saying instrument under leading questioning. Studies that probe VLM perception with direct verbal questioning and no false-alarm baseline are at substantial risk of overestimating perceptual recovery.

### 4.3 Paradigm-recognition confound

Across all 72 reps, the model explicitly named the invisible-gorilla / inattentional-blindness paradigm (often citing Drew, Võ, and Wolfe by name) on 67% of reps, with identical 67% rates on gorilla and control reps. This is a confound with no analogue in Drew’s human study: contemporary LLMs are trained on substantial portions of the cognitive-science literature, including the Simons-Chabris and Drew papers, and they recognize the paradigm by its structural signature (a domain-expert task with an unexpected figure on a final trial).

Three implications follow. First, the elevated control-arm confabulation rate likely reflects an *expectation* of a gorilla derived from paradigm recognition, not a perceptual misclassification of the no-gorilla image. Second, classical cognitive paradigms cannot be applied unmodified to LLM subjects with any guarantee that the model’s response reflects perception of the stimulus rather than recognition of the experimental design. Third, this generalizes beyond inattentional blindness: any extensively described experimental paradigm in the model’s training data is vulnerable to the same confound. Future work on VLMs as cognitive subjects may require either novel paradigms not yet in the literature, or counterbalanced manipulations that isolate paradigm-recognition from stimulus perception.

A “de-Drewed” follow-up experiment isolated the verbal trigger from the stimulus trigger. It reproduced the full Experiment 2 design (three framings × two conditions × 12 reps on gemini-3.1-pro-preview) with a single change: the awareness probes were reworded into neutral phrasing that never names the gorilla and never echoes Drew’s wording. The direct probe, for instance, asked only whether the final case “contained a shape or figure that isn’t part of normal anatomy and doesn’t belong there,” disclosing the category but not the figure. Removing the verbal cues roughly halved explicit paradigm recognition, from 67% to 33% of reps (31% on gorilla reps, 36% on control), confirming that much of the recognition in Experiment 2 had been supplied by the probe language itself. The behavioral signature, however, was unchanged. The model still reported nothing spontaneously during the search task (0 of 24 on both arms); genuine gorilla-versus-control discrimination remained confined to the subtler probes (q2: 50% vs. 8% false alarms, d′ = 1.28); and at the direct and primed levels the gap again collapsed to zero (24/24 vs. 24/24 and 12/12 vs. 12/12, d′ ≈ 0), with control reps confabulating an out-of-place figure, complete with fabricated coordinates, at the same rate as true detections. De-Drewing the probes therefore reduced overt name-checking of the paradigm but did not rescue direct questioning. The residual yes-bias localizes the confabulation to the stimulus rather than the wording: a gorilla rendered in a chest CT is itself the signature of the paradigm, so recognition, and the response bias it produces, persists even when the probe is purposefully neutral. This sharpens the conclusion above, namely that isolating stimulus perception from paradigm recognition requires not merely neutral probe wording but, ultimately, novel stimuli or paradigms not yet represented in the model’s training data.

### 4.4 Dissociation can generalize beyond conversational VLMs

The segmentation-model arm produced a striking cross-over. Both observers — SAM 3 (a generalist trained on natural images) and BiomedParse (a specialist trained on real CT images) — detected the gorilla in its true frame window at ≈92%. But on the gorilla-free control videos, SAM 3 false-alarmed on 0.7% of frames while BiomedParse false-alarmed on 82%. The single-video gorilla-detection rate cannot distinguish a generalist that *localizes the gorilla in time* (SAM 3) from a specialist that *flags “gorilla” everywhere it is prompted to* (BiomedParse). Only the gorilla-free control baseline reveals the cross-over.

This generalization matters in two ways. First, it shows that the methodological lesson of the recreation (that signal-detection theory with an explicit false-alarm baseline is necessary) applies to a model class for which conversational confabulation cannot be the explanation; both segmentation observers produce one-shot per-frame masks rather than verbal responses. Second, the cross-over invites a partial reframing of the human Drew effect: insofar as the radiologists’ inattentional blindness reflected what they had been trained to expect and select for (Most et al., 2005), BiomedParse’s “yes everywhere” pattern is the model-class analogue of an over-attuned specialist, while SAM 3’s narrow precision is the analogue of a generalist for whom the gorilla is the salient anomaly. The mapping is loose, but the false-alarm baseline is what reveals it.

### 4.5 Limitations

Several limitations bear on the inferences drawn here.

#### (i) Statistical power

With n = 12 per cell in Exp 2 (n = 24 pooled across framings), confidence intervals on each probe-level contrast are wide; only the q1 effect survives Holm correction at α = 0.05.

#### (ii) Paradigm recognition

The 67% paradigm-recognition rate means the experiment cannot be characterized as a naive-subject design. We address this by anchoring the central inference on the *signal-detection contrast* (gorilla vs. matched control) rather than absolute YES rates, but the confound cannot be eliminated within this design.

#### (iii) No eye-tracking analogue

We have no measurement of where the model’s processing concentrated during the five trials. Inferences about *where* the model attended rest on response-level dissociations rather than internal-state measurement.

#### (iv) Single-subject deep recreation

Exp 2 is single-VLM (Gemini 3.1 Pro). Experiment 1 partially compensates with breadth (20 model configurations) but at substantially lower stimulus fidelity (static images rather than video stacks). A natural follow-up is the video-stimulus paradigm applied to the full panel.

### 4.6 Implications

For **AI evaluation**, the central methodological takeaway is that any claim of the form “did the model see X” requires an explicit matched-control false-alarm baseline; without one, raw YES rates conflate sensitivity and response bias. This is standard in signal-detection theory but is not yet standard practice in VLM perception benchmarks.

For **cognitive AI**, contemporary VLMs can serve as substrates for testing perception-and-attention theories, but only when the experimental design accommodates two LLM-specific contamination sources: confabulation under leading questioning, and paradigm recognition from training data. Each requires a separate methodological adjustment — control baselining for the first, paradigm-novelty or counterbalanced design for the second. Once both are addressed, the resulting paradigm produces an interpretable, falsifiable signal-detection contrast.

For **radiology AI** specifically, the BiomedParse “yes everywhere” pattern in the segmentation arm is a deployment-relevant failure mode. A medical-image specialist that produces apparent in-window detections at 92% but also false-alarms on gorilla-free controls at 82% would be evaluated as a strong detector under the single-stimulus protocols common in radiology-AI benchmarking. The same model deployed clinically would surface large numbers of false positives at any prompted concept. The point here is not specific to BiomedParse, but to the broader practice of evaluating prompt-conditioned segmentation models without matched-control imagery.

## Data Availability

All data produced in the present study are available upon reasonable request to the authors

## Acknowledgments

This work utilized the computational resources of the NIH HPC Biowulf cluster (http://hpc.nih.gov).

## Data and Code Availability

The analysis code and rep-level JSON result files that support the findings of this study are available upon request of the authors

## Author Contributions

J.D.R. designed and implemented the experiments, performed the analyses, and drafted the manuscript. D.D. and P.H. contributed to study design, methodology, and data analysis. B.D.S. conceived and supervised the study, provided clinical and methodological guidance, and revised the manuscript. All authors reviewed and approved the final manuscript.

## Ethics

This study did not involve human or animal subjects and did not collect new patient data. Existing chest CT imaging was used solely as visual stimuli for evaluating computational models; no patient-identifiable information was accessed or reported.

